# Molecular differentiation by PCR of the *Entamoeba dispar/histolytica/moshkovskii* complex in the population of Coyaima, Tolima, Colombia

**DOI:** 10.64898/2025.12.15.25342303

**Authors:** Juan Ceballos-Castillo, Maria Camila Jurado Guacaneme, Adriana Arévalo, Sonia Dayanni Castillo Ayala, Carlos Franco-Muñoz

**Affiliations:** Universidad Antonio Nariño, Faculty of Sciences, Bogotá D.C., Colombia; Parasitology Group, Public Health Research Division, National Health Institute, Bogotá D.C. 111321, Colombia

**Keywords:** *Entamoeba histolytica*, *Entamoeba dispar*, *Entamoeba moshkovskii*, complejo *Entamoeba*, PCR convencional, Tipificación molecular

## Abstract

The *Entamoeba histolytica/dispar/moshkovskii* complex comprises morphologically indistinguishable species, making accurate diagnosis difficult and leading to overestimations in amebiasis prevalence. This study aimed to establish a sensitive and specific molecular method to differentiate these species in human fecal samples from Coyaima, Tolima, Colombia. A polymerase chain reaction (PCR) targeting the small subunit ribosomal RNA (18S rRNA) gene was developed and optimized under various amplification conditions. The nested PCR showed higher sensitivity and specificity than the multiplex approach, achieving a detection limit of 160 picograms of *E. histolytica* DNA. Cross-reactivity tests confirmed the method’s specificity, as no amplification was observed with other intestinal protozoa or helminths. Epidemiological analysis revealed a prevalence of 24.54% for *E. dispar*, 1.04% for *E. histolytica*, and 0.78% for mixed *E. histolytica/dispar* infections, while *E. moshkovskii* was not detected in the study area. These findings demonstrate the effectiveness of the optimized protocol as a reliable molecular tool for differential diagnosis and epidemiological surveillance of the *E. histolytica/dispar/moshkovskii* complex, contributing to improved understanding of its distribution and transmission dynamics in endemic Colombian populations.

## 1. Introduction

The generous Entamoeba are a group of unicellular protozoa that colonize the digestive tract of a wide variety of vertebrates and invertebrates, of which *E. histolytica* is the one with human health implications causing intestinal and extraintestinal infections, known typically as amoebiasis (1,2). Its life cycle requires the ingestion of the mature cyst (resistant form) form from water of food contaminated, then excystation occurs on the distal ileum and cecum and the trophozoites (active form) colonize the large intestine, then some of the trophozoites are transformed back into cysts and expulsed with the faeces, ensuring their permanence in the environment and perpetuate their biological cycle. (1–3).

Amebiasis caused by *E. histolytica* can range from asymptomatic colonization, mild diarrhea, abdominal pain, to severe clinical presentations, such as amebic colitis, pneumonia, purulent pericarditis and the liver abscesses, this last is the extraintestinal form more common (4,5). This range of presentations may be due to the host’s immune status and the degree of parasite invasion enhanced by *E. histolytica*’s virulence factors (6,7).

Although *E. histolytica* is the pathogenic amoeba, it is morphologically indistinguishable under the microscope with *E. dispar* and *E. moshkovskii* which together form a species complex (8). Amebiasis has a global distribution; its prevalence is higher in regions with limited access to drinking water, basic sanitation systems, and public health education (4).

The estimated prevalence of Entamoeba infection in humans is 3.55%, being the third most common parasitic disease responsible for mortality worldwide (9,10). The highest prevalence is reported in countries such as India, various regions of sub-Saharan Africa, as well as Mexico, Central America, and South America. Recent epidemiological studies have reported a varying prevalence rate 11.1% to 20.2% in Yemen (11,12). And there is confirmed presence of *E. histolytica* in 22/30 countries of the American continent (13).

In 2019, there were 2,539,799 (95% UI 850,865-6,186,972) DALY cases attributable to Entamoeba infection Xiaofang Fu. In Colombia, the National Survey of Intestinal Parasitism in School-Aged Children (2012-2014) revealed that the *E. histolytica/dispar/moshkovskii* complex has a national prevalence of 17%, with the Sierra Nevada of Santa Marta being the most affected region, with a prevalence of up to 48% in this population (14).

Given the limitations of microscopy in differentiating morphologically similar species, other methodologies have been explored, such as *in vitro* culture, isoenzymes and Ag-ELISA which have limitations due to their technical complexity, prolonged processing time and cross-reaction with other Entamoebas (15,16). In this context, Polymerase chain reaction (PCR) has marked a turning point in the diagnosis of amebiasis (17), having now different versions of the technique (nested, multiplex, real time) applicated to a successfully differentiation of *E. histolytica/dispar/moshkovskii* complex with different DNA targets tested, being a laboratory technique catalyzed by the COVID-19 pandemic on the laboratories, allowing the limitations of microscopy and the analysis of non-invasive samples (17–19).

Considering the persistent epidemiological burden of *E. histolytica/dispar/moshkovskii* complex in regions like Colombia, where environmental, social, and health factors coexist that favor transmission (14,20,21), it is necessary to strengthen differential diagnostic approaches (15,22).

The present study aims to optimize and apply molecular tools for the detection and typing of the complex from fecal samples collected in the municipality of Coyaima (Tolima), through the implementation and adjustment of PCR techniques, in order to estimate its regional prevalence and contribute to strengthening the diagnosis and epidemiological surveillance of amebiasis (4,13,17,23).

## 2. Materials and Methods

### 2.1 Study area and population

Coyaima is a municipality located in the department of Tolima, within Colombia’s Andean region (3°47′51″N, 75°11′38″W), at an altitude of 392 m a.s.l. and an average temperature of 28 °C. The municipality comprises 6.20 km^2^ of urban area (1%) housing 18.9% of the population and 658.13 km^2^ of dispersed rural area (99%) where 81.1% of inhabitants live, distributed across 54 microterritories. According to the National Administrative Department of Statistics (DANE), the post-COVID-19 adjusted population in 2024 was 23,363 inhabitants.

This study was conducted within the research project *“Teniasis/Cysticercosis Complex: A Public Health Problem in the Municipality of Coyaima Prioritized in the Andean Region—An Integrated Approach under the One Health Framework”*, funded by the Ministry of Science, Technology, and Innovation (MinCiencias) under Call 918 (2022). The project was executed by the National Institute of Health (INS) in collaboration with the Departmental Government of Tolima, the University of Cauca, and Universidad INCCA de Colombia, with the Ministry of Health and Social Protection as a partner. Ethical approval was granted by the INS Ethics and Research Methodologies Committee (CEMIN, approval No. 14-2021).

As most of the population in Coyaima self-identifies as Indigenous, the study protocol was reviewed and approved by community leaders, including the *Consejo Regional Indígena del Tolima* (CRIT), *Federación de Consejos Autónomos Indígenas del Tolima* (FICAT), and *Asociación de Consejos Indígenas del Tolima* (ACIT). All participants provided informed consent prior to sampling.

From September 2023 to March 2024, a Simple Random Sampling (SRS) design was implemented, following guidelines from the Ministry of Health, based on a registry of 7,602 families in Coyaima. Assuming an expected prevalence of 50%, a 95% confidence level, a 5% margin of error, and an additional 20% to account for potential losses, a target sample of 447 families was established. Randomization was performed in Microsoft Excel, ensuring proportional representation across microterritories. The main food preparer or household head was designated as the sampling unit, from whom fecal and blood samples were collected, along with informed consent and a Knowledge, Attitudes, and Practices (KAP) questionnaire.

Of the 449 participants enrolled, 390 fecal samples were obtained (86.9% of the expected total). Parasitological analyses were performed at the Parasitology Group Laboratory of the National Institute of Health (Bogotá, Colombia) using Kato-Katz and SAF-Ether microscopy. Intestinal parasites were detected in 58% of samples (221/390), with some exhibiting up to five different coinfections. Among pathogenic protozoa, the most frequent were *Entamoeba* complex (29.2%, 114/390) and *Giardia* spp. (3.3%, 13/390). Other detected protozoa included *E. coli* (39.4%), *E. hartmanni* (7.4%), *Endolimax nana* (4.1%), *Iodamoeba butschlii* (3.1%), *Chilomastix mesnili* (1.0%), and *Blastocystis hominis* (19.0%). Helminths detected included *Uncinaria* spp. (51 cases) and *Ascaris lumbricoides* (one case).

### 2.2 Biological Material

A total of 114 fecal samples collected from the municipality of Coyaima, Tolima (Colombia) and previously confirmed as positive for the Entamoeba histolytica/dispar/moshkovskii complex by microscopy were included in this study, out of 383 total samples collected in the region (24). Samples were collected in sterile containers, preserved in absolute molecular-grade ethanol (96–100%), and transported at ambient temperature to the Parasitology Laboratory of the National Institute of Health (INS), Bogotá, for subsequent processing.

Prior to DNA extraction, the fecal material was filtered through a fine mesh to remove coarse debris and facilitate homogenization. One sample was excluded due to insufficient volume, resulting in 113 samples being processed for molecular analysis.

### 2.3 DNA Extraction from Fecal Samples

DNA extraction was performed from ethanol-preserved and pre-filtered fecal samples. Each sample was centrifuged at 3500 rpm for 5 min, and the supernatant was discarded. The resulting pellet was washed three times with 1X phosphate-buffered saline (PBS), repeating centrifugation under the same conditions to remove residual ethanol and particulate matter.

To identify the most efficient extraction protocol yielding DNA of optimal purity and free of PCR inhibitors, three different methods were evaluated using the same clinical fecal sample previously confirmed as positive for *E. dispar*. The protocols compared were as follows:

- **Protocol 1 (Manufacturer’s standard):** Extraction was performed directly following the standard procedure of the QIAamp DNA Stool Mini Kit without any pre-treatment.
- **Protocol 2 (Freeze–thaw cycles):** The washed samples were subjected to one freeze–thaw cycle (−80 °C for 10 min, followed by thawing at room temperature) prior to extraction with the same Qiagen kit according to the manufacturer’s recommendations.
- **Protocol 3 (EDTA incubation):** The washed pellet was resuspended in 0.5 mL of 0.5 M EDTA and incubated at 55 °C for 1 h with constant agitation. DNA extraction was then performed using the QIAamp DNA Stool Mini Kit (Qiagen, Hilden, Germany), following the manufacturer’s instructions. DNA was eluted in 75 μL of pre-warmed

DNA concentration and purity were quantified using a NanoDrop 2000 spectrophotometer (Thermo Fisher Scientific, USA). Absorbance ratios A260/A280 and A260/A230 were used to assess the presence of protein or organic contaminants and to verify the suitability of the DNA for downstream PCR amplification.

### 2.4 Nested Conventional PCR Assays

All PCR reactions were performed under controlled laboratory conditions using physically separated work areas for DNA handling and reaction setup. Each workspace was pre-treated with ultraviolet (UV) light to degrade potential nucleic acid contaminants, and all consumables used were certified as DNase- and RNase-free to ensure template integrity.

The first amplification, targeting the Entamoeba genus, was carried out in a total volume of 20 μL containing 1× PCR buffer, 1.5 mM MgCl_2_, 300 μM of each deoxynucleotide triphosphate (dATP, dTTP, dCTP, and dGTP), 0.063 U/μL of Taq DNA polymerase (Thermo Scientific), and 300 μM of each genus-specific primer: EFv (5′-TAAGATGCACGAGAGCGAAA-3′) and EFr (5′-GTACAAAGGGCAGGGACGTA-3′).

The second amplification, specific for E. histolytica, E. dispar, and E. moshkovskii, was performed under identical reaction conditions in a total volume of 20 μL, incorporating 2 μL of the first-round PCR product as template and 300 μM of each species-specific primer pair:

- E. histolytica: EH1Fw (5′-AAGCATTGTTTCTAGATCTGAG-3′) and EH2Rw (5′-AAGAGGTCTAACCGAAATTAG-3′)
- E. dispar: EdisFw (5′-TCTAATTTCGATTAGAACTCT-3′) and EdisRw (5′- TCCCTACCTATTAGACATAGC-3′)
- E. moshkovskii: EmosFw (5′-GAAACCAAGAGTTTCACAAC-3′) and EmosRw (5′-CAATATAAGGCTTGGATGAT-3′)

PCR amplifications were conducted in a T100™ Thermal Cycler (Bio-Rad). The first-round PCR involved an initial denaturation at 94 °C for 5 min, followed by 30 cycles of denaturation at 94 °C for 1 min, annealing at 56 °C for 1 min, and extension at 72 °C for 1 min, with a final extension at 72 °C for 1 min. The second-round PCR followed the same cycling parameters but consisted of 40 cycles to enhance sensitivity.

The PCR products from the second amplification were analyzed by electrophoresis on 2% (w/v) agarose gels prepared in 1× Tris–Borate–EDTA (TBE) buffer. Gels were stained with GelRed® (Biotium) following the manufacturer’s recommendations. For each reaction, 5 μL of PCR product mixed with loading dye was loaded onto the gel, alongside a 100-1500 bp DNA ladder (PR-G2101 Promega c) as a molecular weight marker. Electrophoresis was performed at 80 V for approximately 1 h.

DNA fragments were visualized under UV light using a Gel Doc™ imaging system (Bio-Rad). The expected amplicon sizes were 439 bp for *E. histolytica*, 174 bp for *E. dispar*, and 553 bp for *E. moshkovskii*.

## 3. Results

### 3.1 DNA Extraction from Fecal Samples

Three DNA extraction protocols—(1) standard commercial procedure, (2) freeze– thaw cycles, and (3) EDTA incubation—were evaluated for efficiency and DNA quality. Significant differences were observed among the methods. The EDTA incubation and commercial protocols yielded the best results, as evidenced by more intense amplification bands in PCR assays (Figure 2A). Although both protocols produced comparable DNA quantities, the EDTA-based extraction was selected for subsequent analyses due to its consistent performance and compatibility with downstream molecular applications.

**Figure 1.**
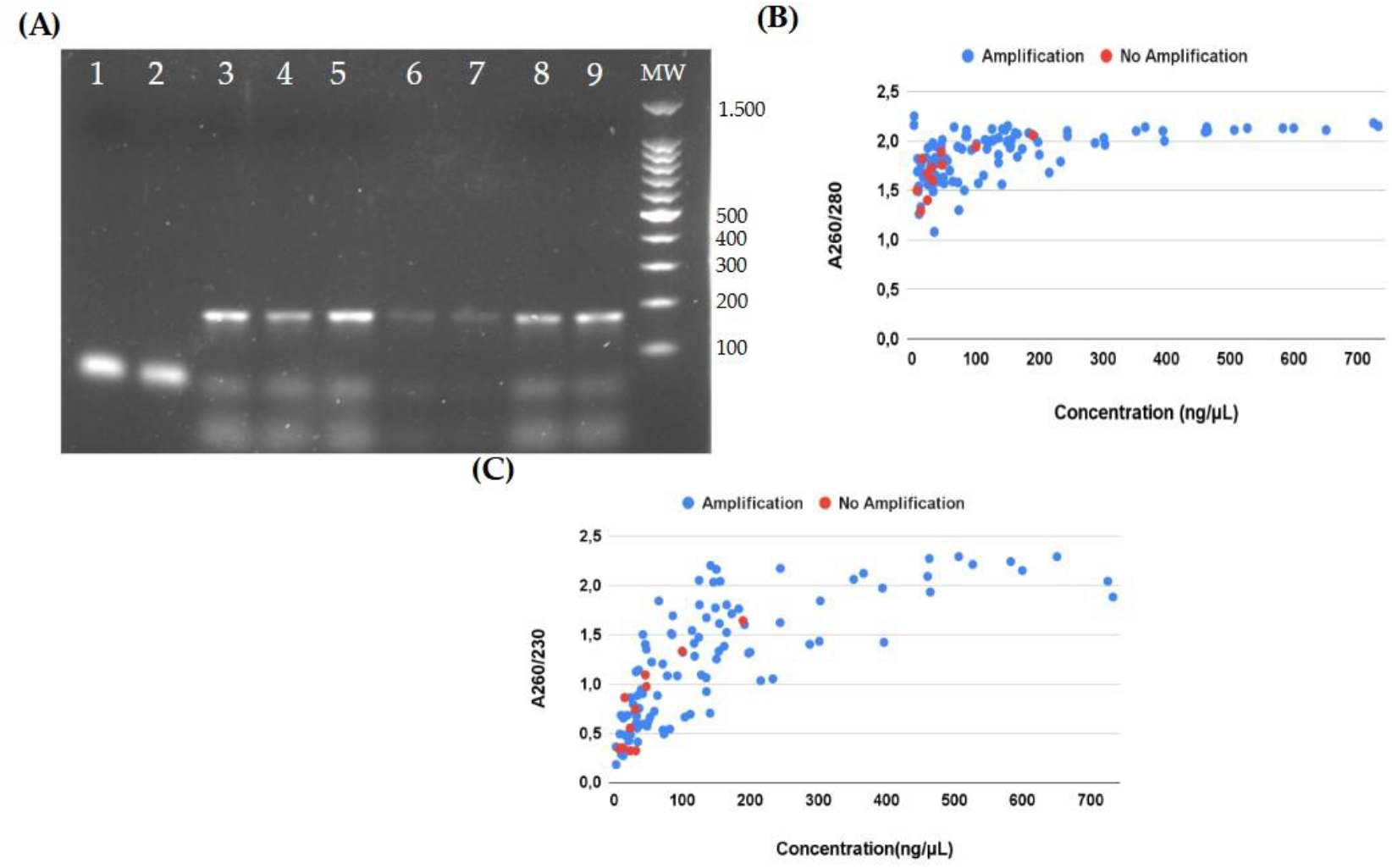
Evaluation of DNA concentration and purity from fecal samples. **(A)** Nested PCR products for the differentiation of the *Entamoeba histolytica/dispar/moshkovskii* complex obtained using three extraction protocols: manufacturer’s standard (no modification), freeze–thaw treatment, and EDTA incubation. Lanes 1 and 2: NTC; lane 3: positive amplification control; lanes 4 and 5: EDTA protocol; lanes 6 and 7: freeze–thaw protocol; lanes 8 and 9: manufacturer’s standard protocol. MW: Promega molecular weight marker (100–1500 bp). (B) DNA concentrations obtained with the selected extraction protocol. (C) A260/280 absorbance ratios and (D) A260/230 ratios of the extracted DNA samples.

**Figure 2.**
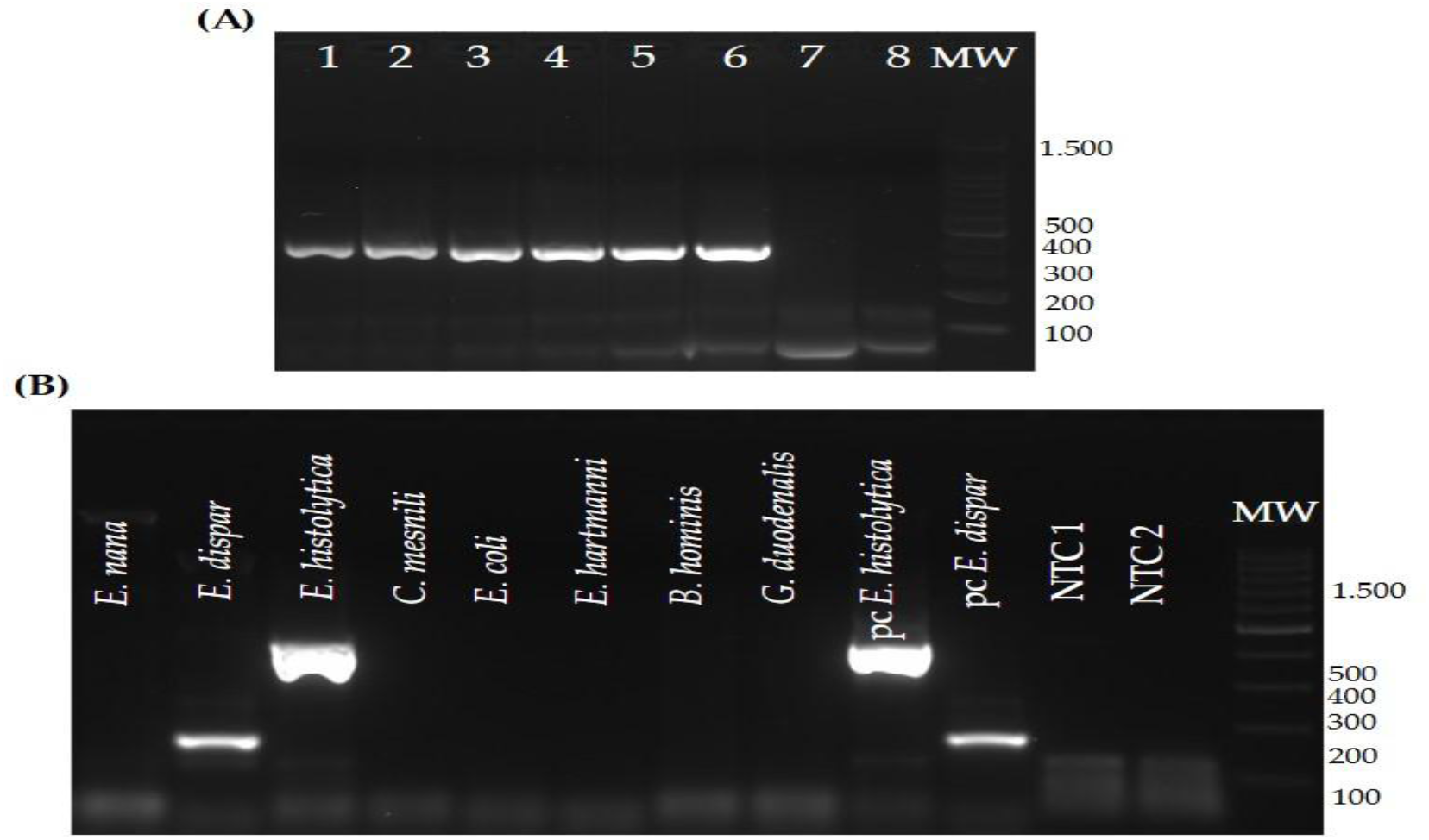
Complementary assays for the optimization of the conventional PCR. **(A)** Limit of detection (LOD) assay of the nested PCR targeting the *Entamoeba histolytica* 18S rRNA gene. Serial fivefold dilutions of genomic DNA were analyzed: lane 1, 100 ng; lane 2, 50 ng; lane 3, 20 ng; lane 4, 4 ng; lane 5, 800 pg; lane 6, 160 ng; lane 7, 32 pg; lane 8, NTC. The expected 439 bp amplicon remained visible up to the lowest concentration tested. MW: molecular weight marker (100–1500 bp); NTC: no-template control. (B) Specificity assay of the nested PCR targeting the *E. histolytica/dispar* 18S rRNA gene. Positive controls (PC) for *E. histolytica* and *E. dispar* and DNA from other intestinal protozoa were tested. Distinct bands of 439 bp and 178 bp correspond to *E. histolytica* and *E. dispar*, respectively. MW: molecular weight marker (100–1500 bp); PC: positive control; NTC: no-template control.

DNA extractions from the 113 ethanol-preserved fecal samples were successful, with concentrations ranging from 2.6 to 732.6 ng/µL. Most samples (79.5%) showed concentrations below 200 ng/µL. A260/280 ratios ranged from 1.08 to 2.25, with higher dispersion observed among samples containing <200 ng/µL DNA (Figure 2B). Ratios <1.7 typically indicate protein contamination or residual extraction reagents such as phenol or guanidine, whereas ratios >2.0 may result from RNA contamination or optical artifacts. Samples with DNA concentrations ≥200 ng/μL exhibited more stable A260/280 ratios (1.9– 2.0), consistent with high-purity DNA suitable for molecular analyses.

A260/230 ratios ranged from 0.18 to 2.29, reflecting variability in sample purity (Figure 2C). Values below 1.8 suggest the presence of common fecal inhibitors such as polysaccharides, bile acids, or residual salts and solvents (25). Despite this variability, no PCR inhibition was observed, confirming that the extracted DNA was suitable for amplification assays. A positive correlation was observed between DNA concentration and A260/230 ratios, indicating that samples with higher DNA yields tended to exhibit improved purity. At concentrations <200 ng/µL, ratios were widely dispersed (0.1–1.5), whereas higher-yield samples (>200 ng/µL) stabilized around 2.0, considered optimal for contaminant-free DNA(26) (25).

Overall, the extraction results demonstrate that, despite the inherent challenges of working with fecal material, the evaluated protocols—particularly the EDTA-based method—produced DNA of sufficient concentration and purity for reliable PCR amplification. These findings align with previous reports highlighting the benefits of pre-purification for improving DNA quality from complex biological matrices (27).

### DNA Extraction from Fecal Samples

An initial multiplex PCR assay was implemented for sample processing, as it is one of the most frequently reported approaches due to its simplicity and low cost. However, experimental assays revealed cross-amplifications between primer pairs, which hindered the clear distinction of specific bands corresponding to *E. dispar* (174 bp), *E. histolytica* (439 bp), and *E. moshkovskii* (553 bp). In addition, low sensitivity was observed in several samples, particularly those with low DNA concentrations. These findings prompted the implementation of a nested PCR system composed of two amplification rounds: a first round targeting the *Entamoeba* genus and a second round specific for *E. histolytica, E. dispar* and *E. moshkovskii*.

The nested PCR markedly improved band resolution and sensitivity, allowing reliable differentiation of positive samples. This approach successfully detected *Entamoeba* DNA in the majority of samples that were negative by multiplex PCR.

During the optimization phase, several parameters were adjusted to enhance amplification efficiency and specificity. MgCl_2_ concentrations ranging from 1.5 to 2.5 mM were tested, with 1.5 mM providing the best balance between yield and specificity by minimizing nonspecific bands. Initial denaturation times of 5 and 10 min were compared, showing no significant differences; therefore, 5 min was selected to reduce total reaction time. Denaturation, annealing, and extension steps were maintained at 30 s each, with standard temperatures of 95 °C and 72 °C for denaturation and extension, respectively.

Annealing temperatures were optimized by gradient testing. For the first-round PCR (genus *Entamoeba*), temperatures from 50 to 58 °C were evaluated, and 51 °C yielded the most consistent results. For the second round (species-specific), a range of 50–62 °C was tested, with 55 °C producing the clearest and most reproducible bands.

Template DNA volumes from 1 to 15 μL were tested. Low volumes (2 μL) were sufficient for samples with high parasite loads but inadequate for low-concentration samples, whereas high volumes (15 μL) caused signal saturation. An intermediate volume of 10 μL provided optimal amplification across samples. For the second PCR, 0.5–8 μL of the first-round product was tested as template; 1.2 μL produced the best reproducibility without nonspecific amplification. The final reaction conditions were established at a total volume of 10 μL, using 2 μL of the first-round product as template.

These optimized conditions improved inter-replicate reproducibility and reduced nonspecific amplification, confirming the suitability of the nested PCR assay for the differential detection of *E. histolytica, E. dispar* and *E. moshkovskii* in fecal samples.

### 3.3 Sensitivity and Specificity of the Optimized PCR Assay

The optimized conventional PCR assay successfully detected genomic DNA from *E. histolytica* at concentrations as low as 160 pg (Figure 2A). Amplification products were clearly visible up to this dilution, confirming the assay’s analytical sensitivity under the established conditions.

Cross-reactivity tests showed no amplification with DNA extracted from fecal samples microscopically confirmed to contain other intestinal protozoa, including *Endolimax nana, Entamoeba coli, Chilomastix mesnili, Entamoeba hartmanni, Blastocystis hominis*, and *Giardia duodenalis* (Figure 2B). Only the expected fragment sizes for *E. histolytica, E. dispar*, and *E. moshkovskii* were detected, indicating specific amplification of the target sequences.

Together, these results demonstrate that the optimized nested PCR assay effectively amplifies low concentrations of *E. histolytica* DNA and selectively discriminates among species of the *Entamoeba* complex without cross-reactivity with other intestinal parasites.

### 3.4 Sensitivity and Specificity of the Optimized PCR Assay

A total of 114 out of 383 fecal samples (29.73%) were positive for the *Entamoeba histolytica/dispar/moshkovskii* complex by microscopy. Using the optimized nested PCR assay, 113 of these samples (99.12%) were successfully genotyped. Based on the total number of samples analyzed, *E. dispar* was detected in 24.54% (94/383), *E. histolytica* in 1.04% (4/383), and mixed *E. histolytica/E. dispar* infections in 0.78% (3/383), while *E. moshkovskii* was not detected. When expressed relative to the microscopy-positive samples (N = 114), *E. dispar, E. histolytica*, and mixed infections accounted for 82.45%, 3.51%, and 2.63%, respectively, whereas 10.6% of samples showed no amplification signal despite being positive by microscopy (Figure 3A).

**Figure 3.**
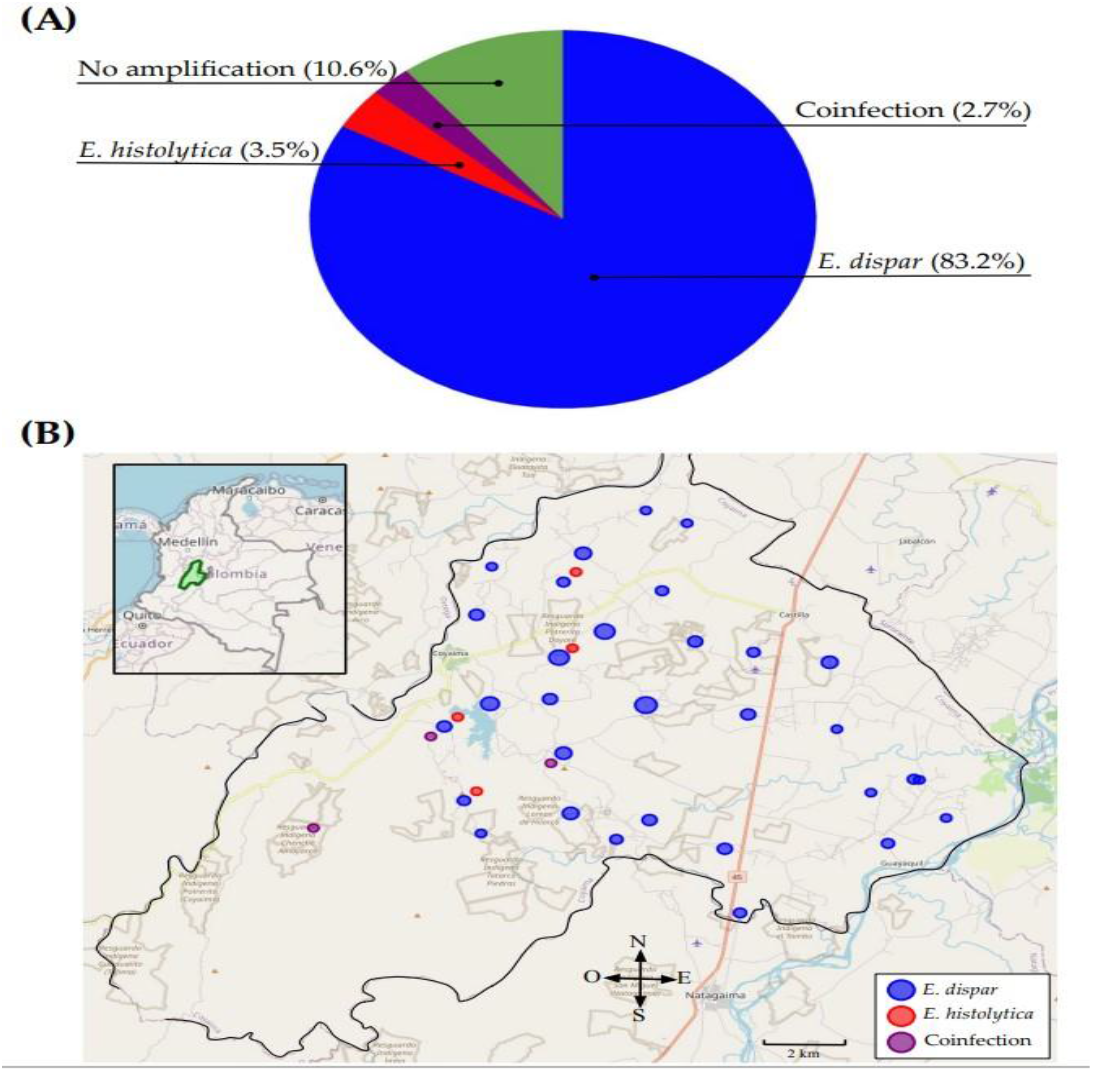
Prevalence and spatial distribution of the *Entamoeba histolytica/dispar/moshkovskii* complex in Coyaima, Tolima. (A) Proportion of *E. histolytica, E. dispar*, and mixed *E. histolytica/E. dispar* infections among microscopy-positive samples for the *Entamoeba* complex (n = 114). (B) Geographic distribution of the *Entamoeba* species detected by nested PCR across the microterritories of the municipality. Map of the Tolima Department created in Python using Folium and GeoPandas, based on the administrative boundaries provided by geoBoundaries (v2.0.0).

The overall prevalence of the *Entamoeba* complex in the study area was 29.2%. Spatial analysis revealed that *E. dispar* was widely distributed across the municipality, whereas *E. histolytica*-positive and mixed infections exhibited a focal distribution concentrated in the western microterritories of Coyaima (Figure 3B). These areas coincide with rural zones characterized by limited access to potable water and basic sanitation, suggesting an environmental influence on parasite transmission dynamics.

Among all samples, 12 (3.13%) identified as positive by microscopy failed to amplify in molecular assays. An endogenous human gene control, assessed by real-time PCR, confirmed amplification in all negative samples. Of the 113 samples confirmed as belonging to the *Entamoeba* complex, 101 yielded detectable amplification, corresponding to an overall PCR positivity rate of 89.38%.

## 4. Discussion

### 4.1 Methodological optimization and DNA quality

The EDTA incubation protocol proved to be the most efficient method for DNA extraction from ethanol-preserved fecal samples, outperforming both the freeze–thaw and unmodified commercial procedures. The superior yield and purity achieved are likely due to the ability of EDTA to chelate divalent cations, inactivate nucleases, and promote cyst disruption, thereby enhancing DNA release. This agrees with previous reports emphasizing the importance of chemical or physical pretreatments to minimize inhibitors in complex fecal matrices (25,27).

The A260/280 and A260/230 absorbance ratios indicated that, although some variability was expected given the nature of the samples, most DNA extracts reached purity levels suitable for PCR amplification, with no evidence of inhibition. These results highlight the effectiveness of the adapted extraction strategy for obtaining high-quality DNA under field conditions, where preservation and transport often represent critical limitations.

### 4.2 PCR optimization and diagnostic performance

The implementation of a nested PCR targeting the 18S rRNA gene successfully overcame the limitations of the multiplex PCR, which included cross-amplification and reduced sensitivity in low-parasite-load samples. The two-step amplification significantly improved sensitivity and specificity, providing sharper and more reproducible bands in agarose gels.

The optimized assay reached a detection limit of 160 pg of genomic DNA, and showed no cross-reactivity with DNA from other common intestinal protozoa. This confirms the fidelity of the selected primers and the robustness of the assay. Collectively, these results validate the proposed protocol as a reliable diagnostic tool for differentiating E. histolytica, E. dispar, and E. moshkovskii, particularly in asymptomatic or low infection contexts.

### 4.3 Molecular epidemiology and spatial distribution of the Entamoeba complex

The overall prevalence of the *Entamoeba histolytica/dispar/moshkovskii* complex in Coyaima (29.2%) falls within the range previously reported for rural and Indigenous communities in Colombia. *E. dispar* was the predominant species (24.54%), followed by *E. histolytica* (1.04%) and mixed *E. histolytica/E. dispar* infections (0.78%), while *E. moshkovskii* was not detected. This pattern corroborates the predominance of *E. dispar* in asymptomatic populations and aligns with national studies reporting its commensal nature and widespread distribution in endemic regions (8,28).

The low frequency of *E. histolytica* observed in Coyaima is consistent with recent evidence of a gradual decline in invasive amebiasis in Colombia, potentially linked to improved access to water and basic health services. In contrast, the absence of *E. moshkovskii* differs from recent reports of its detection in the country (13,29), possibly reflecting demographic or environmental differences among study populations and that it is a recently introduced species.

Spatial analysis revealed that *E. dispar* was widely distributed throughout the municipality, whereas *E. histolytica* and mixed infections displayed a clustered pattern concentrated in the western microterritories of Coyaima. These areas are characterized by limited access to potable water—available only once a week—which may facilitate persistent transmission foci. This spatial trend underscores the relevance of combining molecular tools with geospatial analyses to identify high-risk zones and guide targeted public health interventions in endemic regions.

### 4.5 Technical considerations and limitations

The lack of amplification in 3.13% of microscopy-positive samples was most likely related to low parasite loads or potential misidentification of morphologically similar, non-pathogenic species such as *E. hartmanni, E. polecki*, or *E. coli*. The inclusion of an endogenous human gene control confirmed the integrity of the extracted DNA and ruled out the presence of PCR inhibitors, supporting the reliability of the molecular workflow.

Despite the logistical and economic constraints that limit the routine use of molecular methods in rural settings, this study demonstrates epidemiological value. The establishment of a sensitive, reproducible, and field-applicable assay represents a key step toward strengthening molecular surveillance of amebiasis in vulnerable populations.

### 4.6 Perspectives

The results of this work provide a foundation for developing multiplex PCR systems capable of simultaneously detecting other clinically relevant intestinal protozoa such as *Giardia duodenalis, Cryptosporidium parvum*, and *Blastocystis hominis*, as well as helminths of public health importance. Future studies incorporating quantitative PCR (qPCR) or sequencing approaches could further elucidate the genetic diversity of circulating strains and their relationship to clinical and environmental factors in endemic areas.

## 5. Conclusions

Epidemiological analysis revealed a prevalence of 24.54% for *E. dispar*, 1.04% for *E. histolytica*, and 0.78% for mixed *E. histolytica/dispar* infections, while *E. moshkovskii* was not detected in the study area. These findings demonstrate the effectiveness of the optimized protocol as a reliable molecular tool for differential diagnosis and epidemiological surveillance of the *E. histolytica/dispar/moshkovskii* complex, contributing to improved understanding of its distribution and transmission dynamics in endemic Colombian populations.

## Data Availability

All data produced in the present study are available upon reasonable request to the authors

## Author Contributions

Conceptualization, C.F.-M., A.A., J.C.-C.,. and, M.C.J.G.; Methodology, C.F.-M., A.A., J. C.-C., S.D.C.A. and, M.C.J.G.; Validation, C.F.-M., A.A., J. C.-C., S.D.C.A. and, M.C.J.G; Formal analysis, C.F.-M., J.C.-C. and M.C.J.G.; Investigation, C.F.-M., M.C.J.G., J. C.-C.; writing—original draft preparation, C.F.-M., J.C.-C, M.C.J.G..; writing—review and editing, C.F.- M., M.C.J.G., A.A., J.C.-C. and S.D.C.A.; visualization, J.C.-C. and C.F.-M.; supervision, C.F.-M.; project administration, C.F.-M., M.C.J.G. and A.A. All authors have read and agreed to the published version of the manuscript.

## Informed Consent Statement

Informed consent was obtained from all subjects involved in the study.

## Data Availability Statement

The original contributions presented in this study are included in the article/Supplementary Materials. Further inquiries can be directed to the corresponding author.

## Acknowledgments

To the Ministry of Health and Social Protection, Ministry of Science, Technology, and Innovation, National Institute of Health, Government and Secretary of Health of Tolima, ESE San Roque Municipal Hospital and Coyaima’s mayoralty; to Katerine Gómez Tovar, Yuly Alexandra Loaiza Malambo, and Hermes Jacobo Aguillon Chindoy, who worked in the field; to Coyaima’s community leaders who supported the project; and especially to the community of Coyaima, Tolima.

## References

1. Royer T, Petri WJ. WATERBORNE PARASITES | Entamoeba. 1 de enero de 2014;782–6.

2. Carrero JC, Reyes-López M, Serrano-Luna J, Shibayama M, Unzueta J, León-Sicairos N, et al. Intestinal amoebiasis: 160 years of its first detection and still remains as a health problem in developing countries. Int J Med Microbiol. 1 de enero de 2020;310(1):151358.

3. Soderman WAJ. Microbiología médica [Internet]. 4.^a^. Rama Médica de la Universidad de Texas en Galveston; 1996. Disponible en: https://www.ncbi.nlm.nih.gov/books/NBK7742/

4. Jackson TFHG. Entamoeba histolytica y Entamoeba dispar son especies distintas; evidencia clínica, epidemiológica y serológica. Int J Parasitol. 1 de enero de 1998;28(1):181–6.

5. Ali IKM, Hossain MB, Roy S, Ayeh-Kumi PF, Petri WA, Haque R, et al. Entamoeba moshkovskii Infections in Children in Bangladesh - Volume 9, Number 5—May 2003 - Emerging Infectious Diseases journal - CDC. mayo de 2003 [citado 17 de noviembre de 2025]; Disponible en: https://www.nc.cdc.gov/eid/article/9/5/02-0548_article

6. Carrero JC, Reyes-López M, Serrano-Luna J, Shibayama M, Unzueta J, León-Sicairos N, et al. Intestinal amoebiasis: 160 years of its first detection and still remains as a health problem in developing countries. Int J Med Microbiol IJMM. enero de 2020;310(1):151358.

7. Wilson IW, Weedall GD, Hall N. Host–Parasite interactions in Entamoeba histolytica and Entamoeba dispar: what have we learned from their genomes? Parasite Immunol. febrero de 2012;34(2-3):90–9.

8. López-López P, Martínez-López MC, Boldo-León XM, Hernández-Díaz Y, González-Castro TB, Tovilla-Zárate CA, et al. Detection and differentiation of Entamoeba histolytica and Entamoeba dispar in clinical samples through PCR-denaturing gradient gel electrophoresis. Braz J Med Biol Res. 2019;50:e5997.

9. Cui Z, Li J, Chen Y, Zhang L. Epidemiología molecular, evolución y filogenia de Entamoeba spp. Infect Genet Evol. 1 de noviembre de 2019;75:104018.

10. Singh A, Banerjee T, Khan U, Shukla SK. Epidemiology of clinically relevant Entamoeba spp. (E. histolytica/dispar/moshkovskii/bangladeshi): A cross sectional study from North India. PLoS Negl Trop Dis. 7 de septiembre de 2021;15(9):e0009762.

11. Nath J, Ghosh SK, Singha B, Paul J. Molecular Epidemiology of Amoebiasis: A Cross-Sectional Study among North East Indian Population. PLoS Negl Trop Dis. 3 de diciembre de 2015;9(12):e0004225.

12. Al-Areeqi MA, Sady H, Al-Mekhlafi HM, Anuar TS, Al-Adhroey AH, Atroosh WM, et al. First molecular epidemiology of Entamoeba histolytica, E. dispar and E. moshkovskii infections in Yemen: different species-specific associated risk factors. Trop Med Int Health. 2017;22(4):493–504.

13. Servián A, Helman E, Iglesias M del R, Panti-May JA, Zonta ML, Navone GT. Prevalence of Human Intestinal Entamoeba spp. in the Americas: A Systematic Review and Meta-Analysis, 1990– 2022. Pathogens. noviembre de 2022;11(11):1365.

14. Ministerio de Salud, Universidad de Antioquía. ENCUESTA NACIONAL DE PARASITISMO INTESTINAL EN POBLACIÓN ESCOLARCOLOMBIA, 2012 – 2014 [Internet]. 2015. Disponible en: https://www.minsalud.gov.co/sites/rid/Lists/BibliotecaDigital/RIDE/VS/PP/ET/encuesta-nacional-de-parasitismo-2012-2014.pdf

15. Saidin S, Othman N, Noordin R. Update on laboratory diagnosis of amoebiasis. Eur J Clin Microbiol Infect Dis. 1 de enero de 2019;38(1):15–38.

16. Soares FA, Benitez A do N, Santos BM dos, Loiola SHN, Rosa SL, Nagata WB, et al. A historical review of the techniques of recovery of parasites for their detection in human stools. Rev Soc Bras Med Trop. 2020;53:e20190535.

17. Tawfiq SK, Qader SM, Al-Azzawy MA. Molecular Identification of Entameba Histolytica and Entameba Dispar in Patients with Diarrhea. J Pharm Negat Results. 7 de octubre de 2022;13(4):276–80.

18. Haque R, Kabir M, Noor Z, Rahman SM, Mondal D, Alam F, et al. Diagnosis of Amebic Liver Abscess and Amebic Colitis by Detection of Entamoeba histolytica DNA in Blood, Urine, and Saliva by a Real-Time PCR Assay. J Clin Microbiol. agosto de 2010;48(8):2798–801.

19. Abbas AH, ELShahat ESA, Abdelglil HM. Molecular detection of Entamoeba histolytica in diarrheic fecal samples from patients attending National Hepatology and Tropical Medicine Research Institute. [citado 17 de noviembre de 2025]; Disponible en: https://jmisr.researchcommons.org/home/vol6/iss1/11

20. Espinosa Aranzales AF, Radon K, Froeschl G, Pinzón Rondón ÁM, Delius M. Prevalence and risk factors for intestinal parasitic infections in pregnant women residing in three districts of Bogotá, Colombia. BMC Public Health. 29 de agosto de 2018;18(1):1071.

21. Kann S, Bruennert D, Hansen J, Mendoza GAC, Gonzalez JJC, Quintero CLA, et al. High Prevalence of Intestinal Pathogens in Indigenous in Colombia. J Clin Med. septiembre de 2020;9(9):2786.

22. Fotedar R, Stark D, Beebe N, Marriot D, Ellis J, Jarkness J. Técnicas de diagnóstico de laboratorio para especies de Entamoeba. [citado 17 de noviembre de 2025]; Disponible en: https://journals.asm.org/doi/epub/10.1128/cmr.00004-07

23. Ximénez C, González E, Nieves M, Magaña U, Morán P, Gudiño-Zayas M, et al. Differential expression of pathogenic genes of Entamoeba histolytica vs E. dispar in a model of infection using human liver tissue explants. PLOS ONE. 3 de agosto de 2017;12(8):e0181962.

24. Franco-Muñoz C, Jurado Guacaneme MC, Castillo Ayala SD, Duque-Beltrán S, Arévalo A, Rojas Díaz MP, et al. Baseline Assessment of Taeniasis and Cysticercosis Infections in a High-Priority Region for Taenia solium Control in Colombia. Pathogens. agosto de 2025;14(8):755.

25. Loughrey S, Mannion J, Matlook B. Nucleic acid contamination application ebook [Internet]. Thermo Fisher Scientific, Wilmington, DE; 2022. Disponible en: https://documents.thermofisher.com/TFS-Assets/MSD/Application-Notes/EB53212-acclaro-nucleic-acid-resource-guide.pdf

26. Mohammed F, Taha A, Salama M. Differentiation of Entamoeba histolytica from Entamoeba dispar by nested multiplex polymerase chain reaction. Parasitol United J. 1 de diciembre de 2017;10(1-2):23–9.

27. Verweij JJ, Oostvogel F, Brienen EAT, Nang-Beifubah A, Ziem J, Polderman AM. Short communication: Prevalence of Entamoeba histolytica and Entamoeba dispar in northern Ghana. Trop Med Int Health. 2003;8(12):1153–6.

28. Rivero Z, Villareal L, Bracho Á, Prieto C, Villalobos R. Identificación molecular de Entamoeba histolytica, Entamoeba dispar y Entamoeba moshkovskii en niños con diarrea en Maracaibo, Venezuela. Biomédica. 31 de mayo de 2021;41(Supl. 1):23-34.

29. López MC, León CM, Fonseca J, Reyes P, Moncada L, Olivera MJ, et al. Molecular Epidemiology of Entamoeba: First Description of Entamoeba moshkovskii in a Rural Area from Central Colombia. PLOS ONE. 14 de octubre de 2015;10(10):e0140302.

